# County-Level Proportions of Black and Hispanic populations, and Socioeconomic Characteristics in Association with Confirmed COVID-19 Cases and Deaths in the United States

**DOI:** 10.1101/2020.06.03.20120667

**Authors:** Ahmad Khanijahani

**Affiliations:** Department of Health Administration and Public Health, John G Rangos School of Health Sciences, Duquesne University, Pittsburgh, PA

**Keywords:** COVID-19, Health disparities, Disadvantaged populations, Ethnic and racial minorities

## Abstract

**Objectives:** The objective of this study was to investigate potential county-level disparities among racial/ethnic and economic groups in COVID-19 burden which was measured using confirmed cases and deaths in 100,000 population.

**Design:** Secondary data analysis using county-level data for 3,142 US counties was conducted in 2020. Hierarchical linear regression and concentration curve analyses were performed. The association of COVID-19 cases and deaths was examined separately by sociodemographic and economic characteristics of the county population. American Community Survey (ACS) 5-year estimates (2014-2018), Area Health Resources File (AHRF) 2018-2019, and 2020 COVID-19 data from Johns Hopkins University were used in this study.

**Results:** After adjusting for covariates, US counties with a higher proportion of black population, and a higher proportion of adults with less than high school diploma had disproportionately higher COVID-19 cases and deaths (β>0, p<0.05). A higher proportion of the Hispanic population was associated with higher confirmed cases (β= 1.03, 95% CI= 0.57-1.5), and higher housing cost to household income ratio was associated with higher deaths (β= 3.74, 95% CI= 2.14-5.37). This observed disparities can potentially aggravate the existing health disparities among these population groups.

**Conclusions:** Identification of disproportionately impacted population groups can pave the way towards narrowing the disparity gaps and guide policymakers and stakeholders in designing and implementing population group-specific interventions to mitigate the negative consequences of COVID-19 pandemic.

## Introduction

The Coronavirus Disease 2019 (COVID-19) pandemic has created worldwide challenges for political systems, healthcare systems, public health policy makers, communities, and citizens (Quinn and Kumar 2014). The unprecedented large-scale impact of this highly communicable infectious disease pandemic calls for special attention to disparities in economic and health tolls on different population groups and communities. Health inequities and disparities can be conceptualized and studied with focusing on vulnerable population groups, social determinants of health, and geographical inequalities (Arcaya, Arcaya, and Subramanian 2015; Shi and Stevens 2005; WHO Commission on Social Determinants of Health 2008). Disparities in health status, health outcomes, and access to health services have been shown in different contexts and populations (Williams and Collins 2001; Manuel 2018; Kawachi, Daniels, and Robinson 2005; McQuillan et al. 2004; Lara-Cinisomo, Xue, and Brooks-Gunn 2013). It is well known that concentrated disadvantage plays a significant role in widening the health gap between poor and rich (Browning and Cagney 2002; Jargowsky and Tursi 2015; Braveman et al. 2010). Given the highly contagious nature of COVID-19, the current pandemic demands a population-level understanding of the issue and the consequences. Success in controlling the mortality and morbidity of COVID-19 depends highly on identifying population groups disproportionately impacted by this disease and implementing mitigating policies.

Initial analyses of the individual patient-level data indicate that black and Hispanic minorities are disproportionately hospitalized because of COVID-19 and are more likely to die from the COVID-19 and its complications (Garg 2020; Dyer 2020; Raifman and Raifman 2020). The highly contagious nature of COVI-19 with severe consequences in older populations and those with pre-existing conditions requires behavior in an altruistic fashion with a focus on general welfare rather than self-interested behaviors (similar to the voting behavior studied by Kramer (Kramer)). That is, collective behaviors and aggregated characteristics can potentially be more determinant in the spread of the disease than individual characteristics, supporting the benefit of aggregated county-level analysis.

Using nationwide county-level data from the US, this study aims to examine racial and socioeconomic factors associated with disparities in COVID-19 cases and deaths. It offers preliminary evidence on the existence of county-level disparities on population-size-adjusted COVID-19 cases and deaths to inform policymakers and future studies.

### METHODS

This study considered a total of 3,142 counties in 50 states and the District of Columbia with information available on COVID-19 cases and deaths. Given the ongoing nature of the current pandemic at the time of this analysis (May 18, 2020), 2,837 counties were deemed eligible to be included in the analysis based on a threshold of a minimum of 10 cases in 100,000 population.

Clinical studies show that it takes about 2 to 8 weeks from onset to death in COVID-19 patients (World Health Organization 2020). Because of the ongoing COVID-19 pandemic, the analysis for factors associated with COVID-19 deaths was limited to counties which were reached the epidemic threshold of 10 cases in 100,000 population more than two weeks before the analysis date (on or before May 4, 2020). Thus, 2,738 counties were included in the final analysis of the factors associated with COVID-19 deaths.

#### Data Sources

Table 1 shows the data sources and descriptive statistics for all variables used in this study. To account for differences in county population, all variables are adjusted for county population size and included in the analyses in ratio or percentage forms. The most recent county population estimates for 2019 were obtained from the United States Census Bureau website (United States Census Bureau 2020b)

County-level data for confirmed COVID-19 cases and deaths for all 50 states and the District of Columbia was gathered from the Center for Systems Science and Engineering (CSSE) at Johns Hopkins University (Center for Systems Science and Engineering at Johns Hopkins University 2020). Data was collected from the Centers for Disease Control and Prevention (CDC); local, state and territory health departments; and other sources and are made freely available (Dong, Du, and Gardner 2020)

To ensure the inclusion of all counties with less than 20,000 population and produce the most reliable county-level analysis, American Community Survey (ACS) 5-year estimates (2014-2018), which is based on 60 months of collected data between January 1, 2014, and December 31, 2018, was used (United States Census Bureau 2020a)

The county-level 2018–2019 release of Area Health Resources File (AHRF) was used for several variables. AHRF compiles data from over 50 sources (e.g. American Hospital Association Annual Survey Database, American Medical Association Physician Masterfiles, and Centers for Medicare and Medicaid Services) and provides comprehensive county-level information on topics such as population characteristics and demographics, environment, and distribution of healthcare facilities and professionals (Health Resources and Services Administration 2020)

Four datasets were merged using the county Federal Information Processing Standards (FIPS) code as the unique identifier of each county.

#### Measures

##### COVID-19 Burden

Confirmed COVID-19 cases and deaths in 100,000 population were the outcome variables of interest. To construct these variables, the total number of confirmed cases and deaths in each county until May 18, 2020, were divided by county population and multiplied by 100,000.

##### Disparities

Socioeconomic and demographic variables were used in the original or constructed form to measure disparities. The proportion of Hispanics and black in each county population were used to capture the share of the disadvantaged ethnic and racial combination of the population of each county. National estimates show that about 80% of people living in areas of concentrated poverty are either black or Hispanic (Meade 2014)

Financial hardship was measured in both absolute and relative terms. Median household income (in 2018 inflation-adjusted dollars) was used to capture absolute disparities in county-level differences in income. Household size was controlled in all models. The proportion of households with selected monthly owner cost as a percentage of household income of 35 percent or more (SMOCAPI 35%+, herein) was used to measure relative financial hardship. This variable was constructed by adding the percentage of housing units with or without a mortgage with SMOCAPI 35%+.

##### Covariates

Population characteristics included median age, average household size, percentage female, percentage of population above 25 years old with less than high school diploma, percentage of the civilian noninstitutionalized population with no health insurance coverage, and civilian labor force unemployment rate.

Several variables that can contribute to the spread of COVID-19 and other contagious diseases are included. Population density (persons per square mile) and Core-Based Statistical Area (CBSA) status were included to capture the concentration of the population in each county. CBSA is defined in three categories of Metropolitan, Micropolitan, and Non-CBSA. The percentage of workers 16 years and over who use public transportation (excluding taxicab) to commute to work was included to capture the increased population proximity and temporary concentration of population in enclosed areas.

At the federal level, regardless of other preexisting health conditions or comorbidities, all deaths with confirmed or probable COVID-19 diagnosis are reported as COVID-19 deaths (Centers for Disease Control and Prevention 2020a, 2020b). In an attempt to control for COVID-19 deaths attributable to other causes than COVID-19, all deaths in 100,000 county population from the period of June 2017 to July 2018 was included. Hospital beds per 1,000 population was included to account for the potential differences in the COVID-19 deaths attributable to the hospitalization capacity and availability of hospital personnel.

Four geographical census regions (Northeast, Midwest, South, and West) were included in the analysis to account for the potential impact of environmental factors such as temperature, humidity, and precipitation in the spread of COVID-19 (Pica and Bouvier 2012)

#### Statistical Analysis

To identify county-level factors associated with confirmed COVID-19 cases and deaths, two groups of hierarchical multivariate linear regression models were fit. The two outcome variables were right-skewed and there were more values concentrated around zero. To mitigate the subsequent issues, the outcome variables were transformed to log (confirmed cases in 100,000 population) and log (confirmed deaths in 100,000 population plus one). The unit of analysis was the county. To improve the interpretability of the results, the back-transformed coefficients are reported, which indicate percent changes in the outcome variables.

Cases and deaths reported for counties with smaller denominators (population size) can potentially inflate the ratios disproportionately. Ordering all counties by population size showed that no deaths are reported for counties with a population of less than 1000. Additionally, the least populated counties were not among those with the highest number of cases per 100,000 population.

Besides the variations included at the county level, significant heterogeneity may be present at the state level which can impact both dispersion of cases and the percentage of deaths among the infected persons. To account for differences in magnitude and initiation date of different social distancing actions, along with testing eligibility and reporting criteria and other statewide influential factors, an additional specification was included with state random effects. That is, the models contained a separate level for each state.

Additionally, concentration curves and indices were estimated separately for two outcome variables by plotting the cumulative proportion of the counties ranked by Hispanic ethnicity, black race, median household income, and SMOCAPI 35%+ on the x-axis against the cumulative proportion of cases and deaths on the y-axis. All analyses were performed in Stata MP version 16.1 (StataCorp 2019)

## Results

The characteristics of counties are presented in Table 1. The majority of the variables are measured and reported as a proportion (or percentage) by dividing the county level counts by county population. The county-level average percentage of the population with COVID-19 cases among all 3,142 counties was about 0.25%. The average county-level COVID-19 deaths was about 10 in 100,000 population.

**Table 1.** Characteristics of US Counties, 2020

| Data Source<br>Variable | Mean (SD) | Min | Max |
| --- | --- | --- | --- |
| Johns Hopkins University |  |  |  |
| COVID-19 cases in 100,000 population | 242.14 (521.05) | 0 | 12,256.29 |
| COVID-19 deaths in 100,000 population | 9.91 (23.13) | 0 | 309.83 |
| American Community Survey (ACS) |  |  |  |
| Population | 106,603.3 (360,616.7) | 86 | 10,039,107 |
| % Hispanic | 9.26 (13.79) | 0 | 99.1 |
| % Black | 9.92 (14.7) | 0 | 87.5 |
| Median household income (\$1,000s) | 51.58 (13.7) | 20.19 | 136.27 |
| % SMOCAP 35% or above | 14.73 (4.13) | 0 | 34.93 |
| % 25 years old+ adults with less than high school | 13.41 (6.34) | 1.2 | 66.4 |
| % Female | 49.91 (2.38) | 21 | 58.6 |
| Median age | 41.29 (5.41) | 21.7 | 67 |
| Average household size | 2.52 (0.27) | 1.34 | 4.97 |
| % Uninsured | 10.08 (5.1) | 1.7 | 45.6 |
| % 16-years-old and over commuting to work<br>using public transportation (excluding taxicab) | 0.92 (3.07) | 0 | 61.4 |
| Area Health Resources File (AHRF) |  |  |  |
| Unemployment rate | 4.13 (1.5) | 1.3 | 19.9 |
| Population density per square mile | 259.12 (1724.41) | 0 | 69468.4 |
| Death per 100,000 population (June2017-July2018) | 1,036.06 (267.4) | 0 | 2,181.67 |
| Hospital Beds per 1,000 population | 2.99 (4.92) | 0 | 127.71 |
|  | N (%) |  |  |
| Core Based Statistical Area (CBSA) |  |  |  |
| Not a Statistical Area | 1301 (41.41) |  |  |
| Metropolitan | 1181 (37.59) |  |  |
| Micropolitan | 660 (21.01) |  |  |
| Region |  |  |  |
| Northeast | 217 (6.91) |  |  |
| Midwest | 1055 (33.58) |  |  |
| South | 1422 (45.26) |  |  |
| West | 448 (14.26) |  |  |

**Note.** N= 3,142. COVID-19 cases and deaths are cumulative until May 18, 2020. SMOCAP 35% or above is selected monthly owner costs as a percentage of household income for housing units with or without a mortgage. SD is Standard Deviation.

Figure 1 shows the concentration curves for examining ethnic, racial, and economic disparities in county-level COVID-19 cases and deaths in 100,000 population. The red (45 degrees) line represents the equality line in each graph. In other words, the ranked cumulative percentage on both axes is equal throughout the line. Concerning racial disparities, stronger disparities can be seen in terms of the share of black minorities in the population of the county with both confirmed cases and deaths in 100,000 population (represented by larger shifts from the equality line). Though significant, county-level disparities in Hispanic ethnicity appear to be weaker than racial disparities observed in counties with higher proportions of black populations.

In terms of economic disparities, stronger disparities in both confirmed COVID-19 cases and deaths were observed in counties regarding the SMOCAPI 35%+. The concentration curves with median household income on the x-axis show a higher concentration of population proportionate cases and deaths of COVID-19 in counties with an extremely lower median household income.

**Figure 1.**
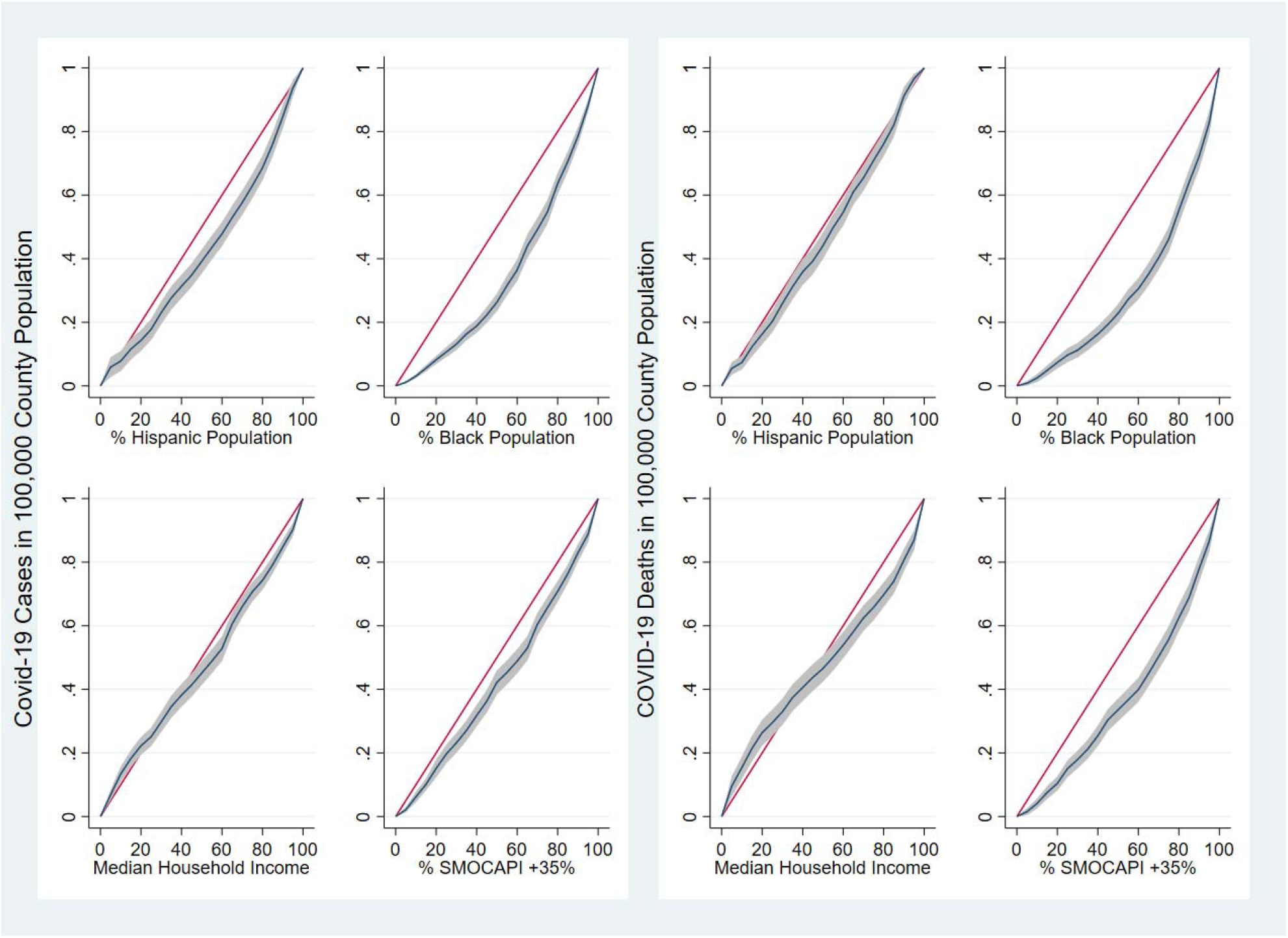
Concentration Curves and Indices Examining County-Level Disparities in COVID-19 Cases and Deaths: the United States, 2020 **Note**. N= 2,837 counties for COVID-19 Cases and 2,738 counties for COVID-19 deaths. All variables in x and y axes are ranked cumulative proportion. The red diagonal line is the line of equity. The grey area around the concentration curve represents 95% confidence interval. SMOCAPI is selected monthly owner costs as a percentage of household income for households with or without a mortgage.

Table 2 shows the results from the multivariate models with COVID-19 cases in 100,000 population as the dependent variable. There were positive relations between the percentage of Hispanic and black populations and COVID-19 cases per population unit. For example, a one-unit increase in the percentage of the black population at the county level was associated with a more than 3% increase in COVID-19 cases in 100,000 (95% confidence interval [CI]: 2.98, 3.7). Higher median income and lower unemployment rate were associated with higher COVID-19 cases per population unit. The second model includes variables for relative financial hardship (SMOCAPI 35%+) and the percentage of 16-year-old or above workers using public transportation (excluding taxicab) to commute to work (commuters, herein). Higher percentage of commuters was associated with higher COVID-19 cases in 100,000 population (β= 0.18, 95% CI= 0.09, 0.28). After including these two variables in the model, the coefficient and significant level for the percentage of Hispanic and black decreased.

**Table 2.**
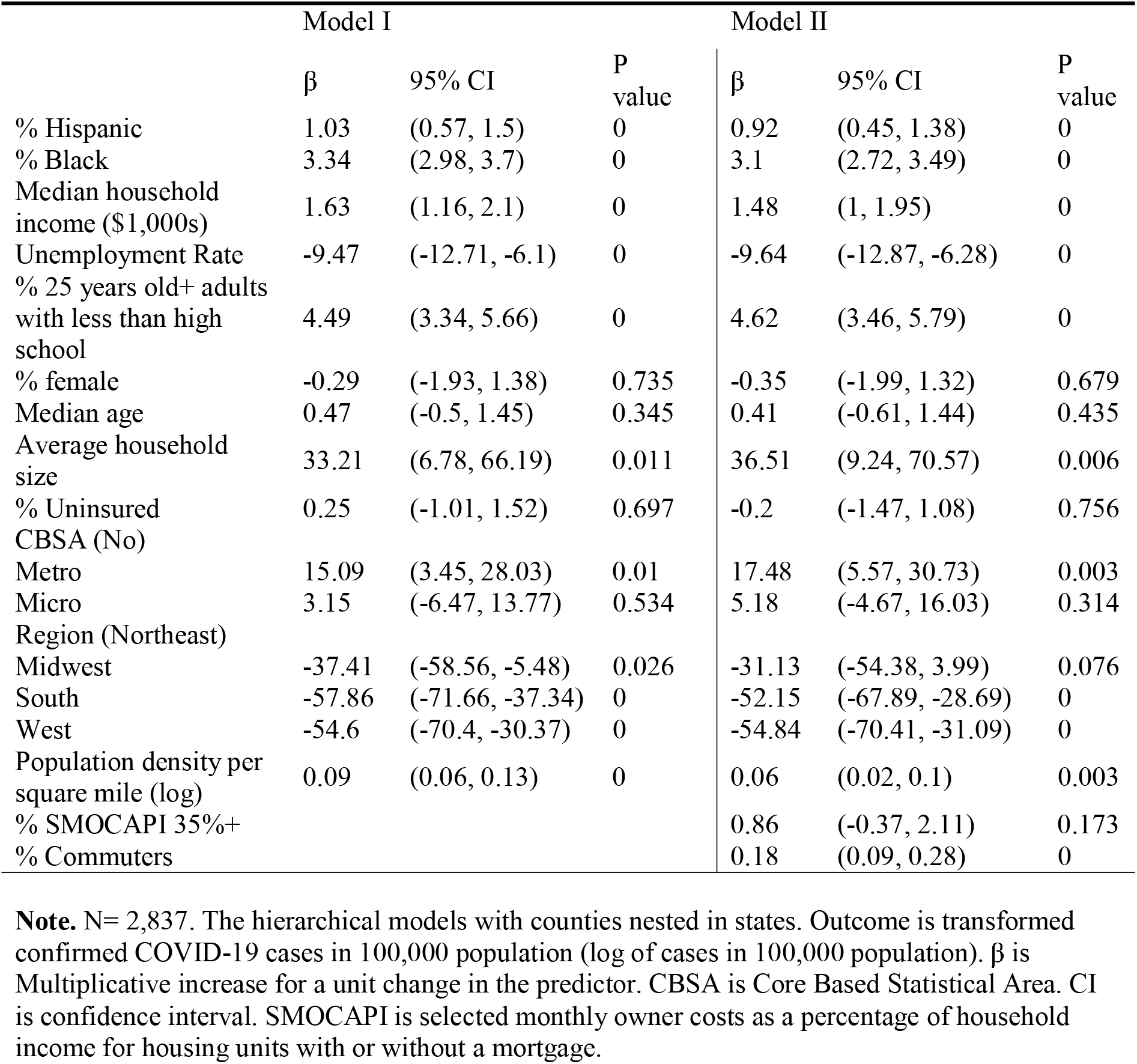
Factors associated with COVID-19 Cases Among U.S. Counties With More Than 10 Cases In 100,000 Population, 2020

Table 3 shows the results from the multivariate models with COVID-19 deaths in 100,000 population as the dependent variable. Positive relations between the percentage of Hispanic and black population and COVID-19 deaths were observed. For example, a one-unit increase in the percentage of black population at the county level was associated with more than 2.5% increase in COVID-19 deaths in 100,000 population (95% CI: 2.08, 3.06). Higher median household income was associated with higher deaths per population unit. COVID-19 cases in 100,000 population in log form was entered into the second model to examine the persistence and change in the coefficients of the predictors. After including this variable in the model, the coefficient for the percentage of Hispanic and black became insignificant (P value>0.1). One percent increase in SMOCAPI 35%+ was associated with more than 3.74 and 2.26 percent increase in per population unit COVID-19 deaths in models with and without COVID-19 cases, respectively.

**Table 3.** Factors associated with COVID-19 Deaths Among U.S. Counties With At Least 10 Cases In 100,000 Population Until 2 Weeks Before the Data Analysis Date, 2020

|  | Model I |  |  | Model II |  |  |
| --- | --- | --- | --- | --- | --- | --- |
| | $\beta$ | 95% CI | P value | $\beta$ | 95% CI | P value |
| % Hispanic | 0.49 | (-0.14, 1.13) | 0.125 | -0.15 | (-0.67, 0.37) | 0.562 |
| % Black | 2.57 | (2.08, 3.06) | 0 | 0.29 | (-0.12, 0.71) | 0.161 |
| Median household income (\$1,000s) | 2.21 | (1.58, 2.84) | 0 | 0.95 | (0.44, 1.47) | 0 |
| Unemployment Rate | -2.01 | (-6.59, 2.79) | 0.405 | 4.89 | (0.82, 9.12) | 0.018 |
| SMOCAPI 35%+ | 3.74 | (2.14, 5.37) | 0 | 2.26 | (0.96, 3.58) | 0.001 |
| % 25 years old+ adults with less than high school | 2.8 | (1.3, 4.33) | 0 | -0.72 | (-1.94, 0.51) | 0.248 |
| % female | 4.38 | (2.15, 6.66) | 0 | 3.91 | (2.08, 5.78) | 0 |
| Median age | -1 | (-2.39, 0.4) | 0.16 | -0.85 | (-2, 0.31) | 0.149 |
| Average household size | 4.3 | (-22.3, 40.02) | 0.779 | -9.09 | (-28.7, 15.9) | 0.442 |
| % Uninsured | -0.4 | (-2.03, 1.27) | 0.64 | -0.25 | (-1.6, 1.12) | 0.72 |
| CBSA (No) |  |  |  |  |  |  |
| Metro | 57.27 | (38.2, 78.97) | 0 | 35.74 | (22, 51.04) | 0 |
| Micro | 29.37 | (14.13, 46.65) | 0 | 24.82 | (12.56, 38.43) | 0 |
| Region (Northeast) |  |  |  |  |  |  |
| Midwest | -45.52 | (-68.46, -5.87) | 0.03 | -24.59 | (-50.35, 14.53) | 0.186 |
| South | -46.12 | (-68.25, -8.57) | 0.022 | -3.6 | (-35.81, 44.78) | 0.86 |
| West | -60.7 | (-77.41, -31.64) | 0.001 | -20.83 | (-48.31, 21.24) | 0.283 |
| Hospital Beds per 1000 population (log+1) | 0.11 | (0.05, 0.18) | 0.001 | 0.09 | (0.03, 0.14) | 0.002 |
| Death per 100,000 population (June2017-July2018) | 0.03 | (0, 0.06) | 0.099 | 0.04 | (0.02, 0.07) | 0.001 |
| COVID-19 cases in 100,000 population (log) |  |  |  | 0.74 | (0.7, 0.78) | 0 |

Note. N= 2,738. The hierarchical models with counties nested in states. Outcome is transformed confirmed COVID-19 deaths in 100,000 population (log of deaths in 100,000 population). $\beta$ is Multiplicative increase for a unit change in the predictor. CI is confidence interval. SMOCAPI is selected monthly owner costs as a percentage of household income for housing units with or without a mortgage.

## Discussion

Using a county-level analytical approach, this study shows how vulnerable ethnic and racial minorities and concentrated financially disadvantaged populations can disproportionately be impacted by COVID-19. According to the findings, a higher percentage of black population in a county was associated with a higher proportion of the population with confirmed COVID-19 cases and deaths. These relations stayed significant even after controlling for other demographic and environmental characteristics, population density, health insurance coverage, and hospital beds, which can impact the spread of contagious diseases and the likelihood of survival or death. Earlier individual-level studies of persons impacted by COVID-19 and consequent deaths indicate the existence of disparities (Dyer 2020; Garg 2020). However, these studies have focused on select groups of patients that might not produce nationally representative samples. Additionally, the county-level examination of the association between concentrated disadvantage and COVID-19 cases and deaths proportion to population highlight the existence and persistence of broader population-level disparities.

Given the highly contagious nature of COVID-19, communities with a higher concentration of vulnerable racial minorities can disproportionately suffer from the global pandemics and regional epidemics of COVID-19 and other infectious diseases. Not only will this issue impact the approximate population, but also it will impede the state-and nation-wide strives to contain and control the spread and burden of COVID-19 and other communicable disease outbreak and epidemics.

A higher share of Hispanic and black ethnic and racial minorities in the population of counties was associated with disproportionately higher ratios of COVID-19 cases and deaths both in regression models and concentration curves and indices. However, counties with a higher proportion of the Hispanic population were more impacted disproportionately by COVID-19 cases than deaths. This can be observed by comparing the coefficients and p values in the regression models and the shift from the equality line along with higher values of concentration indices.

Different factors might contribute to these disparities. For example, Hispanic and blacks are disproportionally living in concentrated disadvantaged communities with higher percentages of the population living below the federal poverty level (Meade 2014). Additionally, compared to their non-Hispanic and white counterparts, percentage of Hispanic and black minorities working in jobs that cannot be fulfilled remotely is disproportionately high (U.S. Bureau of Labor Statistics).

In this study, the impact of county-level economic disparities were measured through the association of median household income and selected monthly owner costs as a percentage of household income of 35% or above (SMOCAPI 35%+) with COVID-19 cases and deaths. Counties in which higher percentages of homeowners spend 35% or more of their household income on housing unit cost were disproportionately impacted by higher COVID-19 deaths. This relation was significant in regression models and concentration curves. Higher housing unit costs can translate to higher rents and can impact both those households who own the housing unit or those who rent. This can be potentially explained in the amount of disposable income left to be allocated to other necessities, savings, and investments. A higher share of income spent on housing might impact the increased COVID-19 cases and deaths in two ways. The lower portion of income left to spend on necessities and lower savings might cause families to need to leave the house more frequently and risk greater exposer to an infectious agent. Additionally, higher housing costs might limit the families’ ability to pay for medical care.

The relation between the median household income and COVID-19 cases and deaths is somewhat puzzling. Although the regression models indicate a positive association between median household income and COVID-19 cases and deaths, the concentration curves depict the relationship with more details. Counties below the about 20^th^ and 40^th^ percentiles of cumulative median household incomes had disproportionately higher cumulative population size-adjusted COVID-19 cases and deaths, respectively. In other words, the counties with the most concentration of the poorest population (those living in extreme poverty) were impacted more severely. This can be observed by a shift of concentration curve to left (above the equality line) in lower percentiles. The Concentration curves for the percentage of families below the Federal poverty level were analogous to the concentration curve for median household income (Results are not reported here).

Besides underscoring the existence of county-level ethnic, racial, and economic disparities in COVID-19 mortalities and mortalities, the findings of this study have several other direct policy implications. Including two variables (SMOCAPI 35%+ and percentage of commuters in county population) in the regression model with COVID-19 cases as the dependent variable, attenuated the contribution of racial disparities and population density. This highlights the importance of financial support and proper social distancing in public transportation in decreasing the speed of the spread of COVID-19. Additionally, by including COVID-19 cases in the regression model with COVID-19 deaths as the dependent variable, the population-level disparities based on the percentage of black and Hispanic populations became insignificant. This suggests that, at least up to this time, disparities in county-level deaths can be mitigated by preventive measures on the dispersion of COVID-19. However, county-level findings should be used by caution for individual person-level interpretations. A future study using a nationally representative sample of COVID-19 patients can clarify the different patterns at the individual level and aggregated county level.

### Limitations

This study had several limitations. First, because all variables in data sources are aggregated and reported at the county level, analysis at an individual level or smaller geographical levels (e.g. census tracts) was not feasible. The aggregation of data for several variables such as gender and age might lead to loss of details, concealment of information, and wrongful inference of ecological level associations to the individual level (Piantadosi, Byar, and Green 1988; Haneuse and Bartell 2011). However, the use of county or other geographical level data is shown to be consistent with individual-lelvel data in several epidemiological and public health studies (McLeod, Nonnemaker, and Call 2004; Marra et al. 2011).

Second, there were several cases and deaths that were not assigned to any county which were only accounted for 0.54% and 1.13% of total confirmed cases and deaths, respectively.

Third, the county-level testing data was not available. Although testing can be a containment measure for controlling the number of cases, at the time of the study, the eligibility for testing was substantially based on the state and local health departments’ and clinicians’ discretion (Centers for Disease Control and Prevention 2020c). Thus, the propensity or ability of a person to pay for the test does not appear to be a major contributor to the number of identified cases.

Finally, clinical studies have found that it takes at least 2 to 8 weeks from onset to death in COVID-19 patients (World Health Organization 2020). Sound inferences about the influencing factors can potentially be made after the end of the current epidemic wave or the passage of enough time from the first within county confirmed COVID-19 case.

## Conclusions

COVID-19 burden appears to vary by aggregated county-level ethnic/racial and economic characteristics of the population. It appears that disproportionate deaths in counties with a higher proportion of the disadvantaged population can be mitigated by proper preventive and social distancing policies accompanied by providing financial support for necessities and medical care.

## Data Availability

The data were derived from the following resources available in the public domain: United States Census Bureau 2020b, Center for Systems Science and Engineering at Johns Hopkins University 2020, United States Census Bureau 2020a, Health Resources and Services Administration 2020.

https://github.com/CSSEGISandData/COVID-19

https://data.hrsa.gov/topics/health-workforce/ahrf

https://www.census.gov/data/tables/time-series/demo/popest/2010s-counties-total.html

https://www.census.gov/programs-surveys/acs

## Data availability

The data were derived from the following sources available in the public domain: United States Census Bureau (2020b), Center for Systems Science and Engineering at Johns Hopkins University (2020), United States Census Bureau (2020a), Health Resources and Services Administration (2020).

## Acknowledgments

Author would like to thank Dr. Christian King and Farzad Aalipour for their comments on the early design of this study. Additionally, author is grateful to Eric Wisz for proofreading this manuscript.

County-Level Proportions of Black and Hispanic populations, and Socioeconomic Characteristics in Association with Confirmed COVID-19 Cases and Deaths in the United States

